# Genetic Risk Factors for Postoperative Atrial Fibrillation – a Nationwide Genome-Wide Association Study (GWAS)

**DOI:** 10.1101/2022.04.29.22274463

**Authors:** Mathias A Christensen, Alexander Bonde, Martin Sillesen

## Abstract

**Background:** Atrial fibrillation (AF) is a major cause of morbidity with a high prevalence among the elderly and has an established genetic disposition. Surgery is a well-known risk factor for AF; however, it is currently not recognized how much common genetic variants influence the postoperative risk. The purpose of this study was to identify Single Nucleotide Polymorphisms associated with postoperative AF.

**Methods:** The UK Biobank was utilized to conduct a Genome-Wide Association Study (GWAS) to identify variants associated with AF after surgery. An initial discovery GWAS was performed in patients that had undergone surgery with subsequent replication in a unique non-surgical cohort. In the surgical cohort, cases were defined as newly diagnosed AF within 30 days after surgery. The threshold for significance was set at 5 × 10^−8^.

**Results:** After quality control, 144,196 surgical patients with 254,068 SNPs were left for analysis. Two variants (rs17042171 (p = 4.86 × 10^−15^) and rs17042081 (p = 7.12 × 10^−15^)) near the *PITX2*-gene reached statistical significance. These variants were replicated in the non-surgical cohort (1.39 × 10^−101^ and 1.27 × 10^−93^, respectively). Several other loci were significantly associated with AF in the non-surgical cohort.

**Conclusion:** In this GWAS-analysis of a large national biobank, we identified 2 variants that were significantly associated with postoperative AF. These variants were subsequently replicated in a unique non-surgical cohort. These findings bring new insight in the genetics of postoperative AF and may help identify at-risk patients and guide management.

## Introduction

Atrial fibrillation (AF) is a widespread morbidity and the most common cardiac arrythmia especially affecting the elderly population.(1) Overall, the prevalence is around 3% rising to 5.9% percent in individuals over the age of 65.(2, 3) The lifetime risk is estimated to be 1 in 4 for men and women older than the age of 40.(4) Besides increasing age, other important risk factors include male gender, congestive heart failure, hypertension, myocardial infarction, diabetes and hyperthyroidism.(5-7)

Genetic risk factors for AF have increasingly been getting attention, and the heritability has been estimated to 22% in the UK Biobank and 62% in a twin study from Denmark.(8, 9) Wolff et al. were the first to describe a family with a hereditary AF trait in *KCNQ1*, coding a K_v_7.1 potassium channel.(10) Since, several other familial loci have been identified, including autosomal dominant traits.(11)

More common genetic variants have since been identified by genome-wide association studies (GWAS), with variants in or near *PITX2* showing the most solid associations.(12) Moreover, genome-wide polygenic risk scores (GPS), calculated on the basis of identified variants from GWAS-models, have managed to identify patients with a three-fold increased risk of AF – a risk that the authors recognize may justify treatment or screening.(13) Postoperative atrial fibrillation (POAF) constitutes its own entity and is the most common secondary form of AF.(14) High rates are especially observed after cardiac surgery, with a prevalence ranging from 20% to 40%, with differences most likely explained by differences in screening rates and monitoring, although factors such as type of procedure, size of the heart and age also may play a major role.(15-18) Even though POAF is often transient, and spontaneous return to sinus rhythm is common, there is still a significant association between POAF after cardiac procedures and increased long-term risk of subsequent persistent AF as well as long-term mortality and risk of stroke.(19-21) POAF is observed at a lower rate after non-cardiac surgery, although the prevalence is still substantial with rates as high as 18%.(22) For non-cardiac surgery, the prevalence is highly variable, most likely due to the heterogeneity of the group, as well as differences in screening protocols, with general rates between 0.5% and 15%.(23-25) Earlier research have sought to identify genetic risk factors for POAF.(26-28) However, these were candidate gene analyses using candidates from previously identified AF risk loci and were only in patients undergoing cardiac surgery.(26-28)

As such, there is currently a lack of studies assessing a potential genetic component of POAF through a standard GWAS approach, as well as to investigate whether genetic variations driving POAF differ from those driving AF in the non-surgical setting. This forms the overall focus of this study.

We hypothesized that common variants may contribute to the risk of 30-day-POAF in a cohort of mixed surgical patients, and that these are overlapping with variants associated with AF in the non-surgical setting.

## Methods

### Phenotypic data

Access to the UK biobank data was approved by the United Kingdom biobank (UKB) consortium (Study ID #60861). Under Danish law, the study was exempt from ethical board approval due to the de-identified nature of the dataset.

We conducted a two-stage GWAS for the risk of POAF. First, a unique cohort of all patients in the UKB with a history of minimally invasive or open surgery was identified. POAF was defined as a new diagnosis (no recorded AF diagnoses prior to surgery) of atrial fibrillation within 30 days after surgery. Subsequently, a second GWAS was conducted in a unique non-surgical cohort to validate variants identified in the surgical cohort, as well as to assess whether an overlap between variants identified in the surgical and non-surgical cohorts could be identified.

UKB is a large, national biobank of more than 500,000 individuals between the age of 40 to 69. All patients were recruited in the years 2006 to 2010 and subsequently followed prospectively for a planned duration of 30 years or longer. All patients currently in the UKB were invited from National Health Service registries and encouraged to complete a comprehensive survey on lifestyle, habits, and medical history. Written consent for participation was given for all participants before inclusion.

### Identification of surgical cohort

Surgical history was identified using OPCS4-codes from UK Biobank data field 41200. Patients with a new diagnosis of AF thirty days post-surgery were classified as cases and those without as controls. POAF was identified using *International Statistical Classification of Diseases, 10*^*th*^ *revision* (ICD-10) and ICD-9 codes. The identifying ICD-10 codes for POAF were I48, I480, I481, I483, I484, I489 and the identifying ICD-9 was 4273. Comorbidities were identified using the appropriate data fields in the UKB and or ICD-10 codes: Heart failure; ICD-10 codes, diabetes; data field 2443.

### Identification of non-surgical cohort

All patients with available genomic data excluding the surgical patients were included for analysis. Cases were defined as patients with a history of AF using the same ICD-10 and ICD-9 codes as defined in the surgical cohort and controls as those without.

By defining each cohort with the above stated criteria no overlapping occurred.

### Quality control of genomic data

Two different arrays were used in the genotyping of patients in UKB. Fifty thousand participants were genotyped using the Affymetrix UK BiLEVE Axiom array while 450,000 were genotyped using the Affymetrix UK Biobank Axiom array. No significant differences exist between the utilized arrays with each genotyping around 850,000 Single Nucleotide Polymorphisms (SNPs).

Quality control was performed using PLINK v1.90b6.16 (Shaun Purcell, MA, US) with a standard approach.(29, 30) Patients with cryptic relatedness, sex discrepancies and outlying heterozygosity rates were excluded. With the purpose of only including patients with high quality genomic information, all individuals with a genotype rate under 98% were excluded. To account for data quality, markers with a missingness rate of more than or equal to 2% were excluded. Further, markers not in Hardy-Weinberg equilibrium were excluded and only variants with a Minor Allele Frequency > 5% were included for analysis.

### Statistics

Both GWAS-analyses in the surgical and non-surgical cohort were analyzed with a mixed linear model approach using fastGWA with a sparse genetic relationship matrix (GRM) using GCTA version 1.93 beta for Windows. A p-value of 5 × 10^−8^ was considered statistically significant. Figures (Manhattan and QQ-plots) were created with qqman (R version 4.0.2).(31) All SNPs are symbolized using the dbSNP Reference (rs) number.

## Results

### Surgical cohort

Overall, 488,377 patients were available in the UBK with genomic data. Following quality control and restriction to surgical patients, 144,196 participants (42,197 excluded from quality control) with 254,068 SNPs were left for analysis. Of these, 1,190 (0.83%) patients had a newly confirmed AF diagnosis up to 30 days after the surgical procedure. Baseline characteristics are listed in table 1.

**Table 1:** Baseline characteristics for discovery surgical GWAS cohort

|  | **Controls** | **Cases** |
| --- | --- | --- |
| Number, N | 143,006 | 1190 |
| Age ± SD, years | 56.5 ± 10.0 | 65.1 ± 6.8 |
| Female, n (%) | 80,270 (56.1) | 336 (28.2) |
| Obesity, n (%) | 9,119 (6.4) | 115 (9.7) |
| Current or past smoker, n (%) | 57,819 (40.4) | 612 (51.4) |
| Major surgery, n (%) | 111,661 (78.1) | 1102 (92.6) |

Two variants reached statistical significance (p < 5 × 10^−8^). The variants were rs17042171 (p = 4.86 × 10^−15^) and rs17042081 (p = 7.12 × 10^−15^). Both SNPs are downstream variants of *PITX2*, a gene previously described in association with AF in other patient populations. The genomic inflation factor, λ, was calculated to be 1.02. The Manhattan plot for this regression is illustrated in figure 1A and the Quantile-Quantile-plot (QQ-plot) is illustrated in figure 2A. A list of the 10 most significant SNPs are listed in table 3.

**Table 2:** Baseline characteristics for replication cohort

|  | **Controls** | **Cases** |
| --- | --- | --- |
| Number, N | 345,103 | 13,205 |
| Age ± SD, years | 55.7 ± 8.1 | 61.9 ± 6.2 |
| Female, n (%) | 188,340 (54.6) | 4,436 (33.6) |
| Obesity, n (%) | 8,660 (2.5) | 1331 (10.1) |
| Current or past smoker, n (%) | 123,230 (35.7) | 6110 (50.2) |

**Table 3:** List of 10 most significant SNPs in the surgical cohort

| SNP | Exon/Intron/Intergenic | Nearest gene | P-value | Associated with AF in non-surgical cohort |
| --- | --- | --- | --- | --- |
| rs17042171 | Intergenic | *PITX2* | $4.86 \times 10^{-15}$ | Yes |
| rs17042081 | Intergenic | *PITX2* | $7.12 \times 10^{-15}$ | Yes |
| rs6740218 | Intergenic | *XPO1* | $3.57 \times 10^{-7}$ | No |
| rs10455872 | Intron | *LPA* | $3.02 \times 10^{-6}$ | No |
| rs56010505 | Intron | *COMMD7* | $3.98 \times 10^{-6}$ | No |
| rs34302613 | Intergenic | *CGNL1* | $4.69 \times 10^{-6}$ | No |
| rs1375302 | Intergenic | *PITX2* | $7.23 \times 10^{-6}$ | Yes |
| rs74617384 | Intron | *LPA* | $7.69 \times 10^{-6}$ | No |
| rs76623075 | Pseudogene | *UCP1* | $7.86 \times 10^{-6}$ | No |
| rs3826653 | Intron | *PPP2R1A* | $9.40 \times 10^{-6}$ | No |

**Figure 1:**
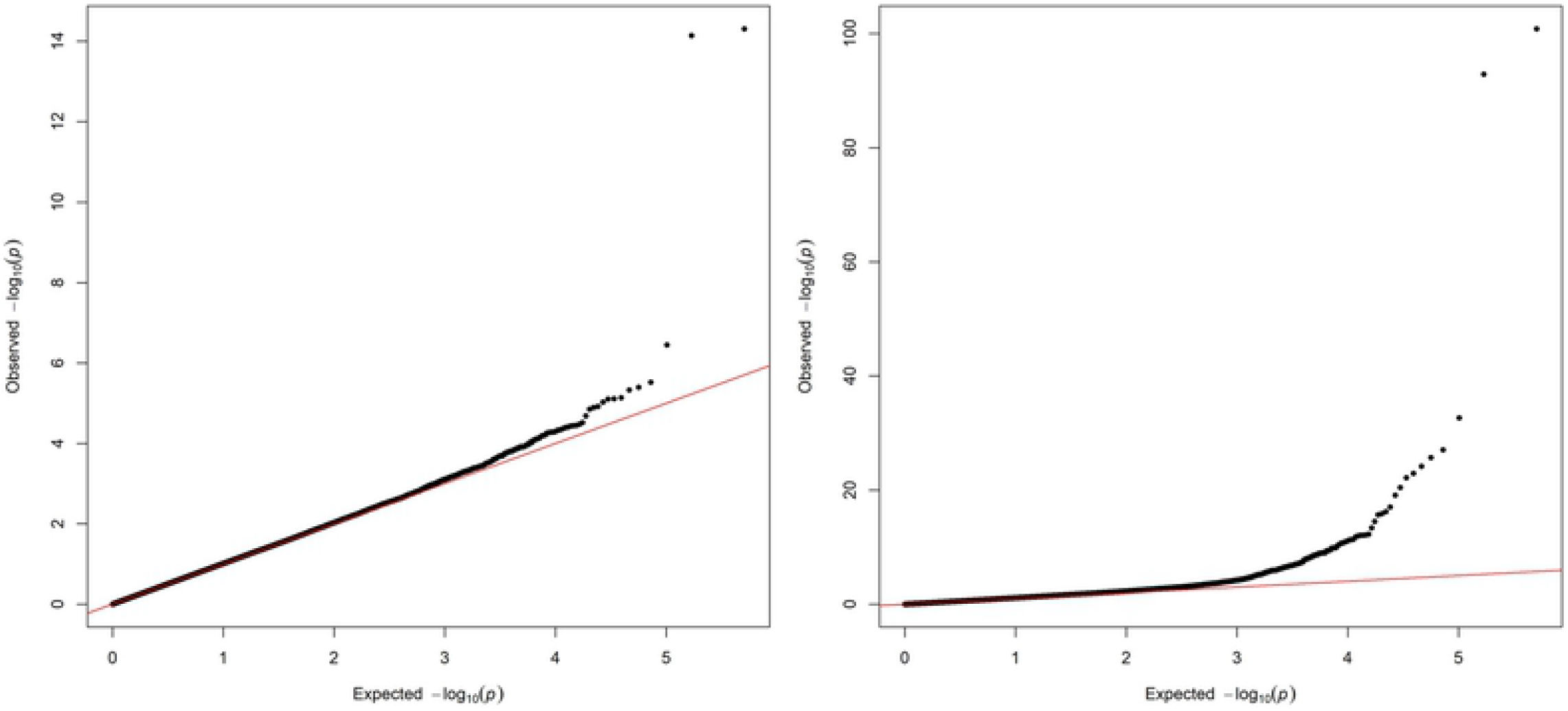
A: Manhattan plot for discovery genome wide association study for new-onset atrial fibrillation 30-days post-surgery Figure 1B: Manhattan plot for replication genome wide association study for atrial fibrillation in the general population

**Figure 2:**
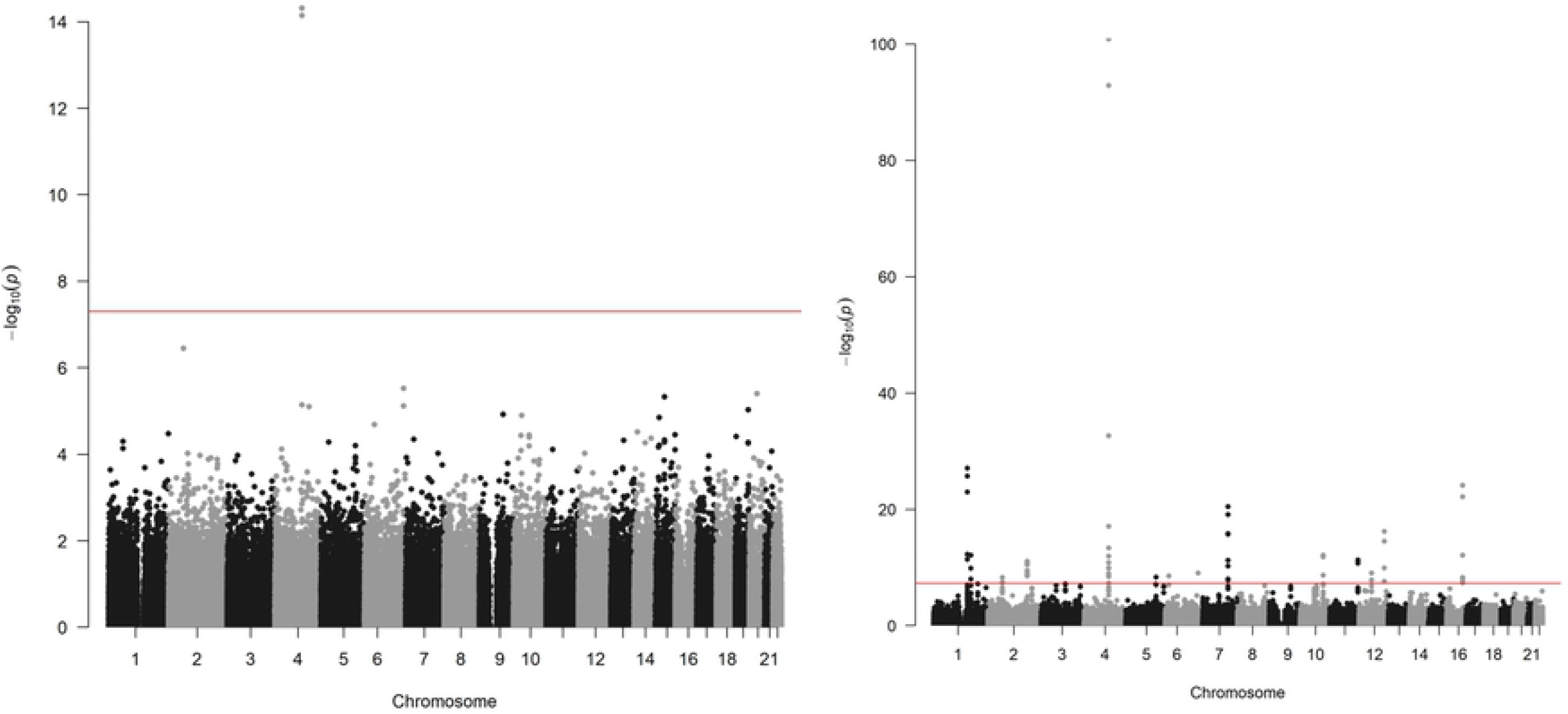
A: Quantile-Quantile plot for discovery genome wide association for new-onset atrial fibrillation 30-days post-surgery Figure 2B: Quantile-Quantile plot for replication genome wide association for atrial fibrillation in the general population

### Non-surgical cohort

Overall, 488,377 patients were available in the UKB with genomic data. Following quality control and exclusion of surgical patients, 301,984 participants (42,192 excluded from quality control) and 254,068 SNPs were left for analysis. Baseline characteristics are listed in table 2. The variants identified in the discovery surgical cohort, rs17042171 and rs17042081, were also significantly associated with AF (1.39 × 10^−101^ and 1.27 × 10^−93^, respectively). Several other loci were found to be associated with AF in this non-surgical cohort, however, none of these loci were found to be associated with POAF. The genomic inflation factor, λ, was calculated to be 1.14. The Manhattan plot for this regression is illustrated in figure 1B and the QQ-plot is illustrated in figure 2B. A list of the 10 most significant SNPs are listed in table 4.

**Table 4:**
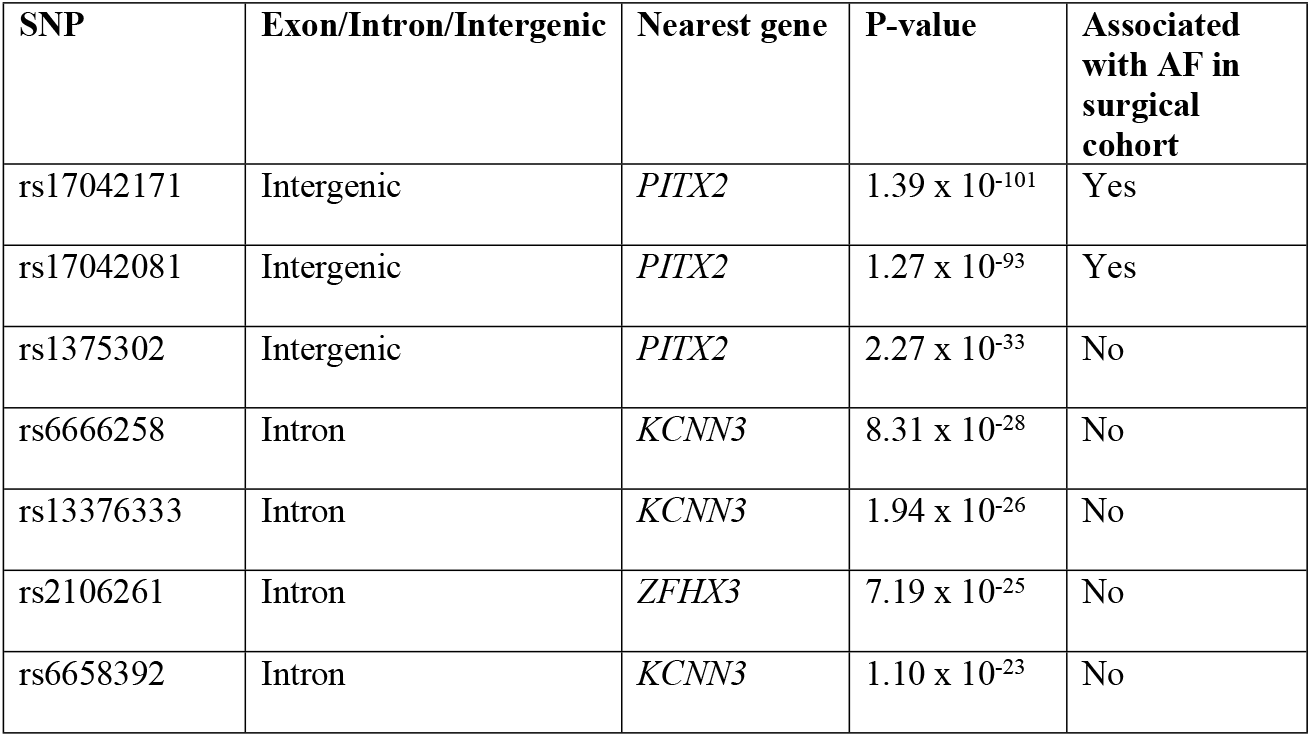
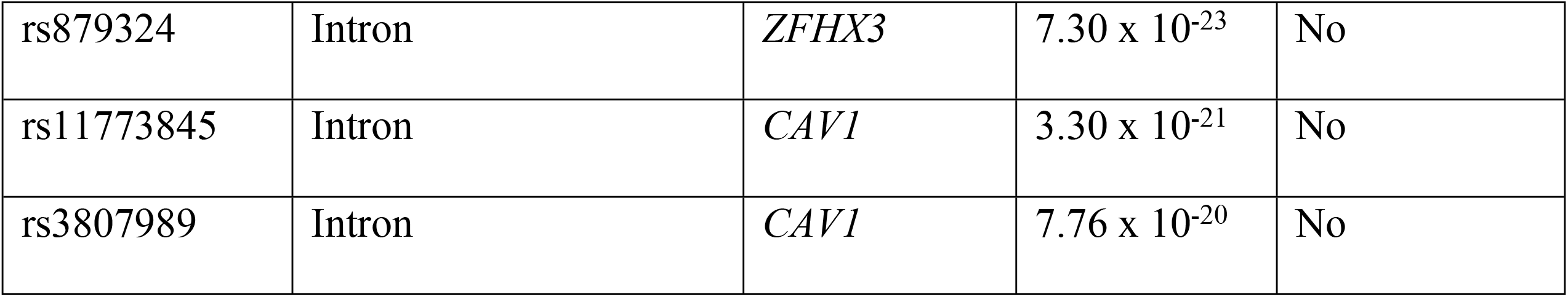
List of 10 most significant SNPs in non-surgical cohort

## Discussion

We describe the findings of a locus associated with 30-day-POAF in a large GWAS-analysis of a surgical population. Further, this locus was replicated and found to be associated with AF in a cohort of non-surgical patients, as well as having been previously associated with AF in other patient populations.(12) Moreover, several loci were found to be associated with AF in the general population without showing any association in the surgical cohort. This could be due to a lack of statistical power but may also be an indicator that fewer genes contribute to the overall risk of POAF compared with general AF, as well as a potential greater contribution of non-genetic factors that we were unable to adequately control for. As some of the pathophysiological mechanisms for POAF differ from general AF, with local inflammation being a contributor to the pathogenesis of POAF, it seems reasonable to assume that the genetic composition of risk factors is also different.(32) However, since POAF is a major risk factor for AF, it is indicative that overlapping factors are also present.(19) The variants found to be associated with both POAF and AF in the general population are downstream variants near *PITX2* on chromosome 4. The gene has previously been associated with AF in various cohorts, including Icelandic, Swedish and Chinese populations.(12, 33) Further, a small candidate gene analysis found an association of the same variants in patients undergoing Coronary Artery Bypass Graft (CABG) Surgery.(26) Additionally, the two variants identified in our study are in very high Linkage Disequilibrium (LD) with the primary risk SNP (rs2200733) identified in this study, which emphasizes the replication of this region as a risk factor. To our knowledge, our analysis is the first large-scale GWAS analysis to describe this association in a heterogenous surgical cohort.

The top variant rs17042171_A near *PITX2* (position: GRCh38.p13: 110787131) is highly prevalent with an overall global frequency of around 16% and a European prevalence of around 13%. The highest prevalence is seen the Asian population, with a prevalence around 50%.(34)

*PITX2* and its genetic product, the paired-like homeodomain transcription factor 2, is a well-described gene and is known to have an essential function in cardiac development including left-right asymmetry, pulmonary vein morphogenesis and shortening of the left atrial action potential.(35-37) The implication in pulmonary vein development is especially interesting as the veins are often the source of ectopic electrical activity involved in the pathogenesis of AF, and often response to radio-frequency ablation.(38) Interestingly, a recent study(39) found that a reduced left atrial expression of *PITX2* and elevated plasma concentrations of the *PITX2*-regulated gene *BMP10* predicted recurrence of AF after catheter-based ablation procedures, further highlighting the role of *PITX2* in the pathogenesis of AF and the importance of large whole-genome studies in identifying potential biomarkers potentially useful in a clinical setting. Moreover, mice studies have shown a promise in *PITX2*-guided pharmacotherapy, as it was indicated that flecainide was superior to sotalol in suppressing AF, explained by *PITX2*-mRNA modulation of atrial membrane resting potential.(37) Although the closest gene to the identified variants is *PITX2*, this analysis does not conclude that *PITX2* is the gene modified by the variants associated with POAF as they are intergenic. However, given the role of *PITX2* in cardiogenesis and the other well-described effects on AF modulation, and the fact that other nearby genes are microRNA and non-functional, it is deemed the most plausible. Moreover, this analysis cannot conclude that the variants identified are the functional modifiers of transcription, as other non-identified SNPs in LD may be.

Interestingly, variants near *PITX2* were the only significant locus in the surgical cohort. Meanwhile, several loci were associated with AF in the general population. Other interesting loci associated with AF in this cohort include rs6666258, an intron variant in *KCNN3*, which encodes a potassium channel protein which is important for atrial repolarization and has previously been associated with AF in other patient populations(40), as well as the very common rs11773845, an intron variant in *CAV1* encoding caveolin-1 which has a great variety of functions and has also been associated with AF in other patient populations.(41) While a lower power may explain the lack of significance of these loci in the surgical cohort, it is also possible that *PITX2* contribute to a greater proportion of the genetic risk of POAF compared with general AF. This is further emphasized by the fact that none of the significant loci from the non-surgical cohort had a suggestive p-value of less than 5 × 10^−5^, which is generally considered the cut-off.(42) However, we utilized a large cohort, and with a disease prevalence of around 3%, an allele relative risk of 1.5, 1,190 cases and around 140,000 controls, a statistical power of > 80% would be expected. With these conditions, it would be expected that our analysis would miss common variants with minor contribution to the overall genetic risk, again suggesting that variants near *PITX2* may be more important in the POAF phenotype compared with general AF.

Genotyping for AF or POAF is not currently common in clinical practice, except for rare familial clusters. Given the morbidity associated with POAF a future approach may include genotyping in preoperative risk assessment, however, this analysis does not currently present an indication. First, replication needs to be done in an independent surgical cohort, secondly, it is currently unknown if genotyping for POAF will decrease the associated morbidity and mortality.

### Limitations

This analysis has limitations. First, this is a retrospective case-control study, and hence comes with the inherent limitations, including the inability to assess causation.

Secondly, the diagnoses of AF and POAF were made with ICD-9 and ICD-10 codes which may be inaccurate and underreported. Asymptomatic cases are not reported, and cases of POAF may have had AF before the procedure, but not given the diagnosis until after, and therefore not as a direct result of the surgery. Consequently, if any true differences in the genomic risk factors for AF and POAF exist, this may have been influenced by this limitation. Further, the data lacked information on postoperative medication, which may confound the results.

Lastly, the study may have been underpowered to detect variants with minor contributions to the overall genetic risk. Our power calculation assumed a relative risk of 1.5 of the variants in question, however, to detect variants with a relative risk of 1.1 more than 20,000 cases would be needed for a power of 80%.

## Conclusion

In conclusion, we report the association of intergenic variants downstream of *PITX2* with postoperative atrial fibrillation in a GWAS-study of a large, national cohort. This finding was replicated in a unique non-surgical cohort with AF. Further, variants associated with AF in the general population were not associated with POAF, indicating that the genomic landscape of AF and POAF may differ.

## Data Availability

There are restrictions prohibiting the sharing of data in this manuscript. The data were obtained from UK Biobank upon application. Access can be applied for at the UK Biobank directly, at http://www.ukbiobank.ac.uk. Acceptance of application will give access to the same data as used for this study.

## Acknowledgements

Data is not publicly available but can be applied for at https://www.ukbiobank.ac.uk/enable-your-research/apply-for-access. Analytic methods will be made public at github.com at request.

## Author contributions

Conceptualization: MC, AB and MS. Data Curation: MC, AB. Formal Analysis: MC, AB. Funding Acquisition: MS. Methodology: MC, AB and MS. Resources: MS. Software: MC and AB. Supervision: AB and MS. Visualization: MC and AB. Writing – Original Draft Preparation: MC. Writing – Review & Editing: AB and MS

